# Prior immune checkpoint inhibitor exposure, treatment interval, and severe hepatotoxicity with sotorasib in KRAS G12C-mutant non-small-cell lung cancer: a systematic review and meta-analysis

**DOI:** 10.64898/2026.10.05.26364739

**Authors:** Seungho Cook, Han Sang Kim

## Abstract

**Background:** Severe hepatotoxicity has been reported when sotorasib follows immune checkpoint inhibitor (ICI) therapy, and fixed washout intervals have been proposed. We assessed prior ICI exposure and the ICI-to-sotorasib interval as separate risk factors.

**Materials and methods:** We searched PubMed, Europe PMC, Embase, Web of Science, Scopus, CENTRAL, and meeting proceedings for comparative studies of grade 3 or higher hepatotoxicity after KRAS G12C inhibitors. Matched risk ratios (RRs) and unadjusted cohort data were synthesized separately, the latter with an exact conditional odds ratio (OR). Certainty was rated with GRADE.

**Results:** Of 530 unique records screened, seven sources were included, all involving sotorasib. In the only matched analysis (meeting abstract, 386 patients per arm), prior ICI exposure was associated with higher risk (13.2% vs 4.4%, RR 3.00, 95% CI 1.76–5.10), whereas matched comparisons at 6 weeks (RR 1.11, 0.61–2.03) and 12 weeks (RR 1.08, 0.52–2.27) detected no difference but could not exclude important effects. All five unadjusted cohorts showed higher risk with shorter intervals, including 21/74 versus 8/75 patients in the earlier versus later interval quartiles of the CodeBreaK 200 trial, although effect sizes were heterogeneous (exact homogeneity P=0.006). Certainty was very low throughout.

**Conclusions:** Prior ICI exposure was associated with higher risk of severe sotorasib-associated hepatotoxicity, with very low certainty. A shorter interval was associated with higher risk in unadjusted data only, and current evidence does not establish a mandatory washout period. Liver-test monitoring per the prescribing information applies to all patients, whereas additional surveillance after ICI exposure remains an untested precaution.

**HIGHLIGHTS:**

- Prior ICI exposure was linked to a threefold higher risk of severe sotorasib hepatotoxicity in a matched analysis.
- Shorter ICI-to-sotorasib intervals carried higher risk in all five unadjusted cohorts, including a phase 3 trial.
- Matched comparisons at 6 and 12 weeks showed no interval effect, so a fixed washout is not established.
- Exact conditional methods handled zero-event comparator arms without continuity correction.
- All comparative evidence was of very low certainty, and adjusted prospective data are needed.

## INTRODUCTION

KRAS G12C is a targetable driver alteration in advanced non-small cell lung cancer (NSCLC), and covalent KRAS G12C inhibitors are established options after first-line therapy.^1–3^ Because first-line treatment usually contains an anti-PD-1 or anti-PD-L1 antibody, most patients who start a KRAS G12C inhibitor have recently received an immune checkpoint inhibitor (ICI).

Severe liver injury after sotorasib started soon after ICI therapy was first described in case reports, including biopsy findings resembling immune-mediated hepatitis.^4^ Checkpoint antibodies persist for months after the last infusion, and similar sequence-dependent hepatotoxicity has been described with other targeted agents given after ICIs.^5–7^ Retrospective cohorts and a meeting abstract subsequently reported high rates of grade 3 or higher hepatotoxicity when sotorasib followed recent anti-PD-(L)1 therapy,^8–12^ and a pooled analysis of sotorasib trials reported more any-grade hepatotoxicity after recent immunotherapy.^13^ These reports led to proposals of washout periods ranging from 30 days to 6 weeks or longer.^8,9^

Two questions are often conflated. The first is whether prior ICI exposure itself identifies patients at higher risk. The second is whether the interval between the last ICI dose and sotorasib initiation modifies that risk, which is the premise of a fixed washout. The existing studies differ in design, comparator, outcome definition, and interval cut-off, and most comparisons are unadjusted with zero events in the comparator group. A propensity-score-matched analysis presented in 2026 reported higher risk after prior ICI exposure but did not detect an interval-associated difference at 6 or 12 weeks.^14^

We therefore conducted a systematic review and meta-analysis that keeps adjusted and unadjusted evidence in separate layers, uses methods appropriate for sparse data, and addresses prior exposure and interval as distinct questions.

## MATERIALS AND METHODS

### Protocol and reporting

The review follows PRISMA 2020 and, for the components synthesized without meta-analysis, the Synthesis Without Meta-analysis (SWiM) guideline.^15,16^ The review was registered in OSF Registries on 4 October 2026 (doi:10.17605/OSF.IO/X3R9Q).

### Eligibility criteria

Eligible studies included adults with advanced KRAS G12C-mutant NSCLC treated with any KRAS G12C inhibitor, and reported grade 3 or higher hepatotoxicity (aminotransferase, alkaline phosphatase, gamma-glutamyl transferase, or bilirubin increase, or drug-induced liver injury) according to prior ICI exposure or ICI-to-inhibitor interval, with a comparator group. Cohort studies, trial subgroup or appendix analyses, and registry analyses were eligible. Full articles, meeting abstracts, and preprints were eligible. We excluded case reports, series without a comparator, studies reporting only any-grade hepatotoxicity or composite toxicity outcomes, pharmacovigilance disproportionality analyses, and trials of concurrent ICI plus KRAS G12C inhibitor combinations. The last three categories are described narratively.

### Information sources and search

PubMed and Europe PMC were searched on 26 September 2026. The Europe PMC search was rerun on 4 October 2026, and Embase, Web of Science Core Collection, Scopus, and the Cochrane Central Register of Controlled Trials were searched on 4 and 5 October 2026 through the Yonsei University library. Proceedings of the American Society of Clinical Oncology, European Society for Medical Oncology, and the International Association for the Study of Lung Cancer World Conference on Lung Cancer (2021 to 2026) were searched through journal supplements. Reference lists of included studies, citing articles, and trial appendices were screened. Full search strings for every source are given in Supplementary Table S1.

### Study selection and data extraction

Screening, full-text assessment, and data extraction were performed with AI assistance (see the declaration on generative AI), and each extracted value was verified independently by the authors against the source document. Disagreements were resolved by re-examination of the source and, when needed, by the senior author. For every extracted number, the exact supporting sentence or table cell was recorded, and each number was labelled as reported or derived. Derived values were limited to arithmetic combinations of reported categories. Event counts were not reconstructed from rounded percentages.

We extracted design, setting, drug, exposure definition, comparator, interval categories, hepatotoxicity definition and grading, event counts, and adjusted effect estimates. For interval comparisons, we used each study’s available split nearest to 3 months (12 weeks, 90 days, or 3 months), and for CodeBreaK 200 the first two versus the last two reported interval quartiles around a rounded 2.60-month boundary. The finest reported categories were extracted for descriptive display.

### Risk of bias and certainty of evidence

Risk of bias was assessed at the domain level with ROBINS-E, because prior ICI therapy and its timing were not randomized in any study.^17^ Judgments were made at the domain level, because most sources did not report enough detail to answer every signalling question. Certainty of evidence was rated with GRADE separately for each question and evidence layer.^18^

### Synthesis methods

Evidence was synthesized in two layers. The adjusted layer contained reported propensity-score-matched RRs, which were presented as reported and were not converted or combined with unadjusted data. The unadjusted layer contained study-level 2×2 tables.

Because most comparator arms had zero events, the unadjusted layer was pooled with an exact conditional (Fisher noncentral hypergeometric) common OR, conditioning on the margins of each table, which requires no continuity correction.^19–21^ The conditional maximum likelihood estimate is reported when finite, with an exact 95% CI and a two-sided exact P value based on probability ordering. When every comparator arm has zero events the upper bound is not identified, and only the exact lower bound is reported. Heterogeneity was assessed with an exact conditional likelihood-ratio test of homogeneity of ORs, following Zelen’s conditional framework,^22^ and with the asymptotic Breslow–Day test with Tarone correction. A conditional random-effects model and Bayesian random-effects models with six prior combinations were fitted as supplementary analyses. Sensitivity analyses excluded the trial appendix data or the meeting abstract by Kagalwalla et al. and omitted each study in turn. Absolute risk differences with Newcombe CIs are reported per study.

Studies whose comparator was not an ICI-naive or longer-interval group (Chour et al., who compared ICI in the immediately preceding line with a mixed control) were described separately. Analyses used Python with custom exact conditional routines, which were implemented with AI assistance and checked independently by the authors. Code and extraction files are available in the OSF project (see Data availability). Small-study effects and reporting bias were not assessed statistically because fewer than ten studies contributed to each comparison. Their likely influence was considered in the GRADE ratings.

## RESULTS

### Study selection

Database searches retrieved 648 records (PubMed 25, Europe PMC 413, Embase 92, Scopus 88, Web of Science 20, CENTRAL 10). After removal of 118 duplicates, 530 records were screened, and 18 reports were assessed in full. Three further reports were identified through meeting proceedings, reference lists, and trial appendices (Figure 1). Full-text exclusions are listed in Supplementary Table S2. Seven sources met the eligibility criteria (Figure 1). Six contributed quantitative data to the two synthesis layers, and one (Chour et al.) was described separately. The principal full-text exclusions were a large national cohort whose ICI-stratified comparison covered all grade 3–4 treatment-related adverse events rather than hepatotoxicity,^23^ a veterans cohort reporting a composite of dose modifications for diarrhea, hepatotoxicity, or pneumonitis,^24^ and a pooled trial analysis reporting only any-grade hepatotoxicity.^13^ In the Italian expanded access programme, 23 of 26 previously treated patients with grade 3–4 treatment-related adverse events had received prior anti-PD-(L)1 therapy, but hepatotoxicity was not reported by ICI exposure (Supplementary Table S2). A previously cited interval comparison attributed to Chour et al. (16/44 vs 6/51) could not be found in the original report, the published commentary, or the authors’ response, and was removed.

**Figure 1.**
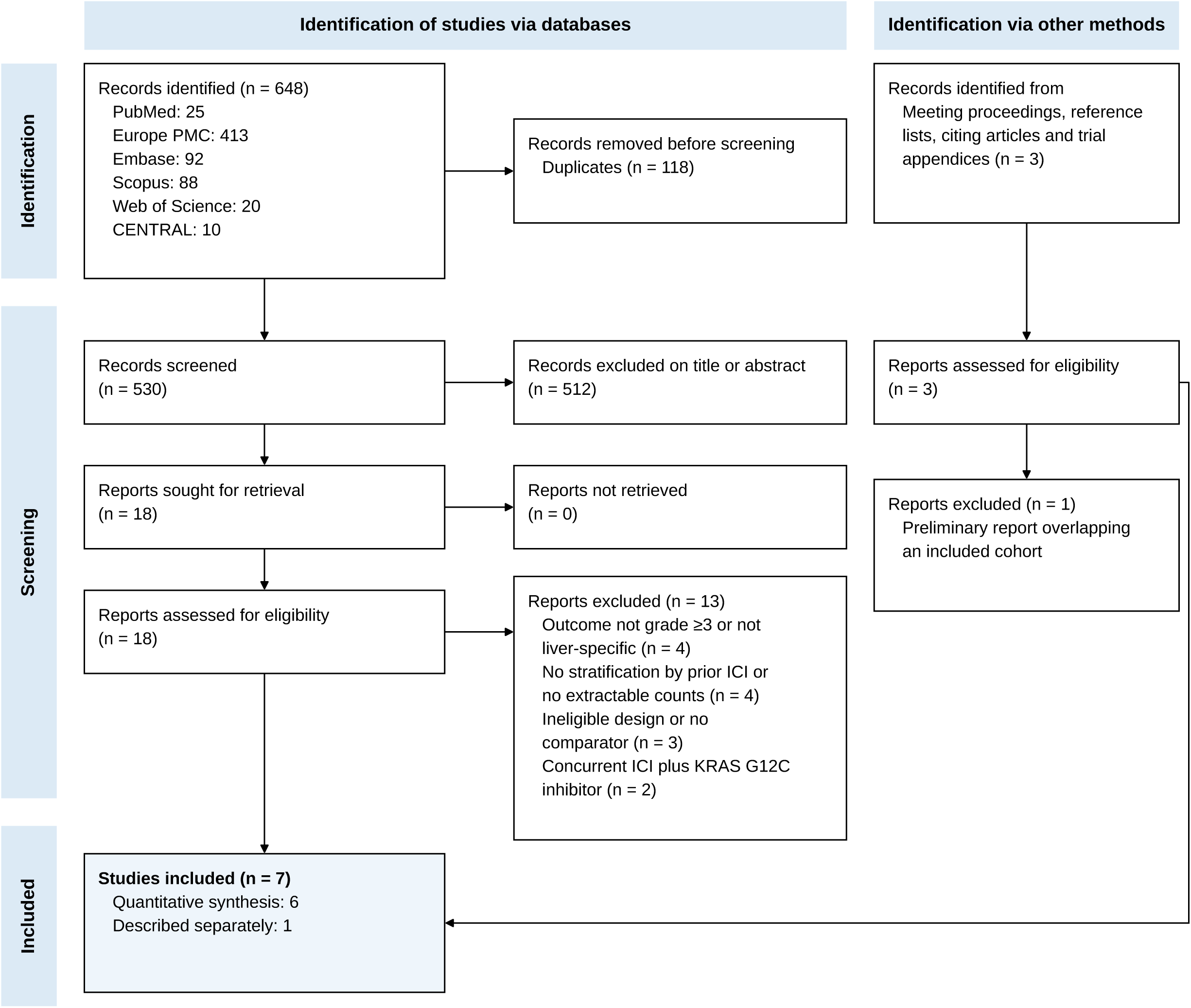
PRISMA 2020 flow diagram. Duplicates were identified by normalized title matching across databases. Exclusion reasons for reports assessed in full are listed in Supplementary Table S2.

### Study characteristics

Characteristics of the included sources are summarized in Table 1. All involved sotorasib. No eligible comparative data were identified for adagrasib or other KRAS G12C inhibitors. The matched analysis by Patel et al. was a retrospective real-world study reported as a meeting abstract, with 1:1 propensity-score matching on age, race, sex, liver metastasis, alcohol-related disorders, liver disease, obesity, and statin use.^14^ Its outcome was labelled grade 3 or higher hepatotoxicity and was operationally defined as aspartate or alanine aminotransferase more than three times the upper limit of normal or total bilirubin above 3 mg/dL within 90 days. The abstract lists 458 patients per arm in its Methods but reports 386 per arm after matching, without describing the stage of the Methods sample or the reasons for the difference.

**Table 1.** Characteristics of included sources.

| Study | Design and setting | Exposure contrasts | Hepatotoxicity definition | Data, events/N (reported unless stated) | Synthesis role |
| --- | --- | --- | --- | --- | --- |
| Patel 2026<sup>14</sup> | Retrospective real-world cohort, 1:1 propensity-score matching. Meeting abstract, database and sites not reported | ICI within 1 year vs ICI-naïve<br><6 vs ≥6 weeks<br><12 vs ≥12 weeks | AST/ALT >3×ULN or total bilirubin >3 mg/dL within 90 days (labelled grade ≥3) | 13.2% vs 4.4% (386/arm)<br>11.7% vs 10.5% (171/arm)<br>11.5% vs 10.6% (113/arm)<br>Event counts not reported | Adjusted layer |
| Ernst 2024<sup>9</sup> | Prospective biomarker cohort (NCT05221372), single centre, Netherlands | Prior anti-PD-(L)1 vs none<br>≤6, 6–12, ≥12 weeks | CTCAE v5 grade ≥3 ALT, AST, ALP, or GGT increase | 28/80 vs 0/11<br>15/18, 8/24, 5/38 (≤6 and 6–12 vs ≥12 weeks: 23/42 vs 5/38, derived) | Unadjusted layer |
| Desai 2023<sup>11</sup> | Retrospective, Mayo Clinic sites, USA | Prior ICI vs none<br>≤30, 31–90, >90 days | Grade ≥3 hepatotoxicity | 10/28 vs 0/3 (derived from 10/31 overall, 28 with prior ICI, 0/3 ICI-naïve)<br>3/4, 7/11, 0/13 (≤90 vs >90 days: 10/15 vs 0/13, derived) | Unadjusted layer |
| Thummalapalli 2023<sup>10</sup> | Retrospective multicentre, USA | Prior anti-PD-(L)1 vs none<br>≤12 vs >12 weeks | Grade ≥3 sotorasib-related hepatotoxicity | 13/86 vs 0/19 (derived from text and Table 3)<br>13/53 vs 0/33 (derived from linked interval and liver-specific statements) | Unadjusted layer |
| Kagalwalla 2024<sup>12</sup> | Retrospective, single centre; meeting abstract | Prior ICI vs none<br><3 vs ≥3 months | CTCAE v5 grade 3–4 transaminitis | 4/23 vs 0/5 (derived)<br>4/16 vs 0/7 (0/7 derived from grade 1–2-only events in that group) | Unadjusted layer |
| CodeBreak 200, de Langen 2023<sup>2</sup> | Phase 3 trial, sotorasib arm; appendix Table S16 (post hoc subgroups) | Prior immunotherapy vs none<br>Quartiles of time since last immunotherapy (<1.58, 1.58–2.60, 2.60–6.21, >6.21 months) | CTCAE v5 grade ≥3 hepatic events of interest (Hepatic Disorders SMQ, narrow); not restricted to treatment-related | 29/149 vs 3/20<br>12/36, 9/38, 5/36, 3/39 (quartiles 1–2 vs 3–4: 21/74 vs 8/75, derived) | Unadjusted layer |
| Chour 2023<sup>8</sup> | Retrospective, 16 French centres | Anti-PD-(L)1 in the immediately preceding line vs mixed control (87% earlier ICI with intervening regimen) | CTCAE v5 grade ≥3 sotorasib-related hepatotoxicity | 16/48 vs 6/54 | Described separately |

All sources involved sotorasib. Derived values are arithmetic combinations of reported categories. ALP, alkaline phosphatase; ALT, alanine aminotransferase; AST, aspartate aminotransferase; CTCAE, Common Terminology Criteria for Adverse Events; GGT, gamma-glutamyl transferase; ICI, immune checkpoint inhibitor; SMQ, standardised MedDRA query; ULN, upper limit of normal.

The unadjusted layer comprised four clinical cohorts and the sotorasib arm of the CodeBreaK 200 phase 3 trial.^2,9–12^ In CodeBreaK 200, appendix Table S16 reports grade 3 or higher hepatotoxicity events of interest (Hepatic Disorders standardized MedDRA query, narrow) by prior immunotherapy and by quartile of time since the last immunotherapy dose. Table S16 lists treatment-related adverse events and grade 3 or higher hepatic events of interest separately, and it does not establish that all hepatic events of interest were attributed to treatment by investigators. This was the only source with events in the ICI-naive group.

### Prior ICI exposure

In the matched analysis, ICI exposure within one year before sotorasib was associated with a higher risk of hepatotoxicity than ICI-naive status (13.2% vs 4.4%, RR 3.00, 95% CI 1.76–5.10, P<0.001; Figure 3A).^14^

In the unadjusted layer, 84 of 366 ICI-exposed patients and 3 of 58 ICI-naive patients across five cohorts had severe hepatotoxicity (Table 2, Figure 2A). Four cohorts had no events among ICI-naive patients. The pooled exact conditional OR was 5.14 (95% CI 1.59–26.45, P=0.002). The exact homogeneity test gave P=0.059 (Breslow–Day with Tarone correction, P=0.158). Without CodeBreaK 200, the conditional maximum likelihood estimate and upper confidence limit were unbounded (exact 95% CI 2.87 to infinity). Within CodeBreaK 200 alone, severe hepatic events occurred in 29 of 149 (19.5%) patients with and 3 of 20 (15.0%) without prior immunotherapy (OR 1.37, 0.36–7.76).

**Figure 2.**
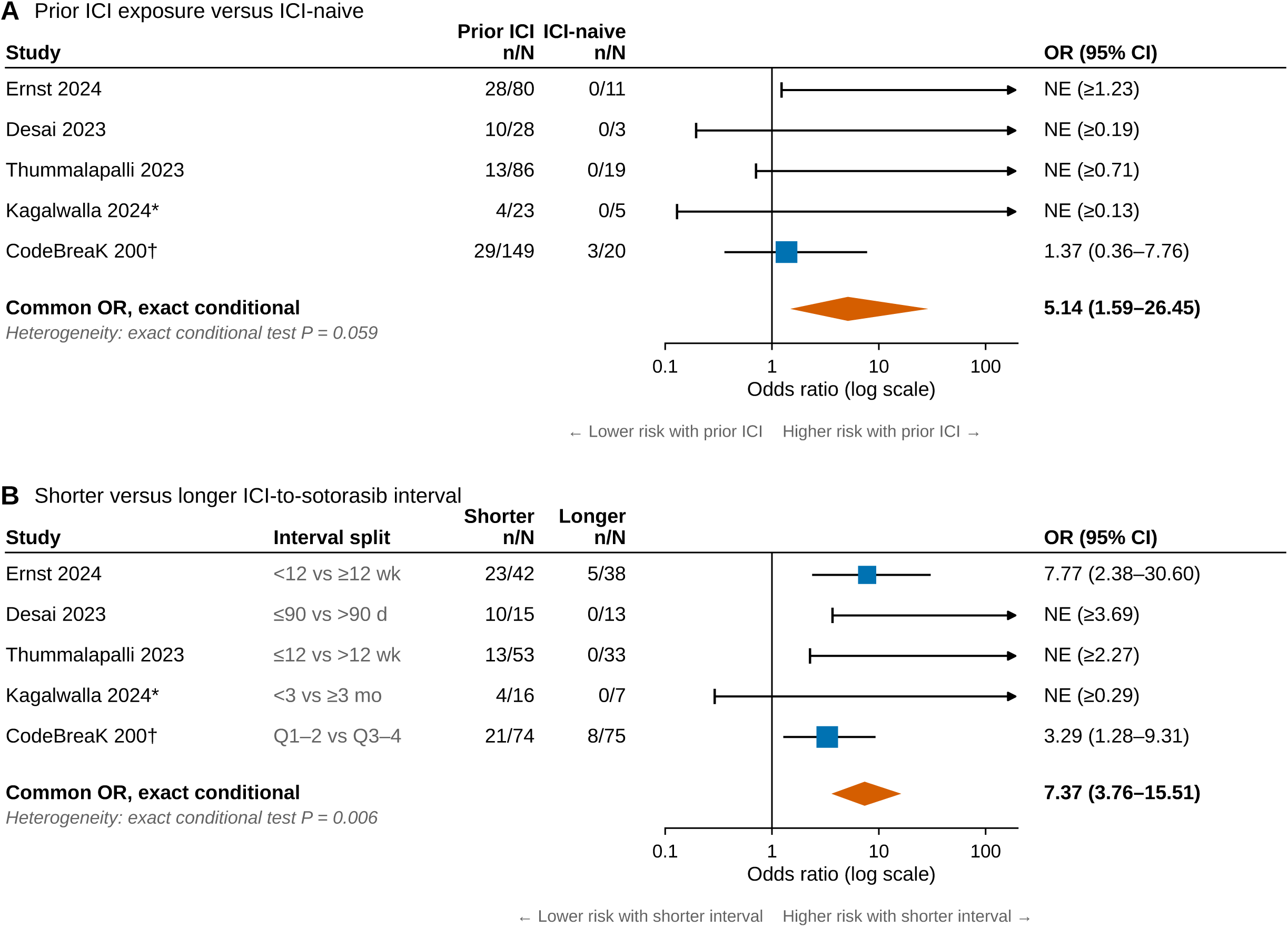
Unadjusted cohorts, exact conditional analysis. (A) Prior immune checkpoint inhibitor (ICI) exposure versus ICI-naive status. (B) Shorter versus longer interval between the last ICI dose and sotorasib, using each study’s split closest to 12 weeks. Squares show study-level conditional maximum likelihood odds ratios (ORs), with size proportional to study sample size. Where the comparator group had no events, the OR is not estimable (NE), the exact lower 95% confidence limit is marked, and the arrow indicates an unbounded upper limit. Diamonds show the exact conditional common OR. Without CodeBreaK 200, the common OR was not estimable for panel A (exact 95% CI 2.87 to infinity) and was 15.18 (5.42–53.44) for panel B. *Conference abstract. †Phase 3 trial appendix (Table S16), grade 3 or higher hepatic events of interest (Hepatic Disorders standardized MedDRA query, narrow), not restricted to treatment-related events. Pooled estimates assume a common OR, are not adjusted for confounding, and are not combined with the matched risk ratios in Figure 3. CI, confidence interval.

**Table 2.**
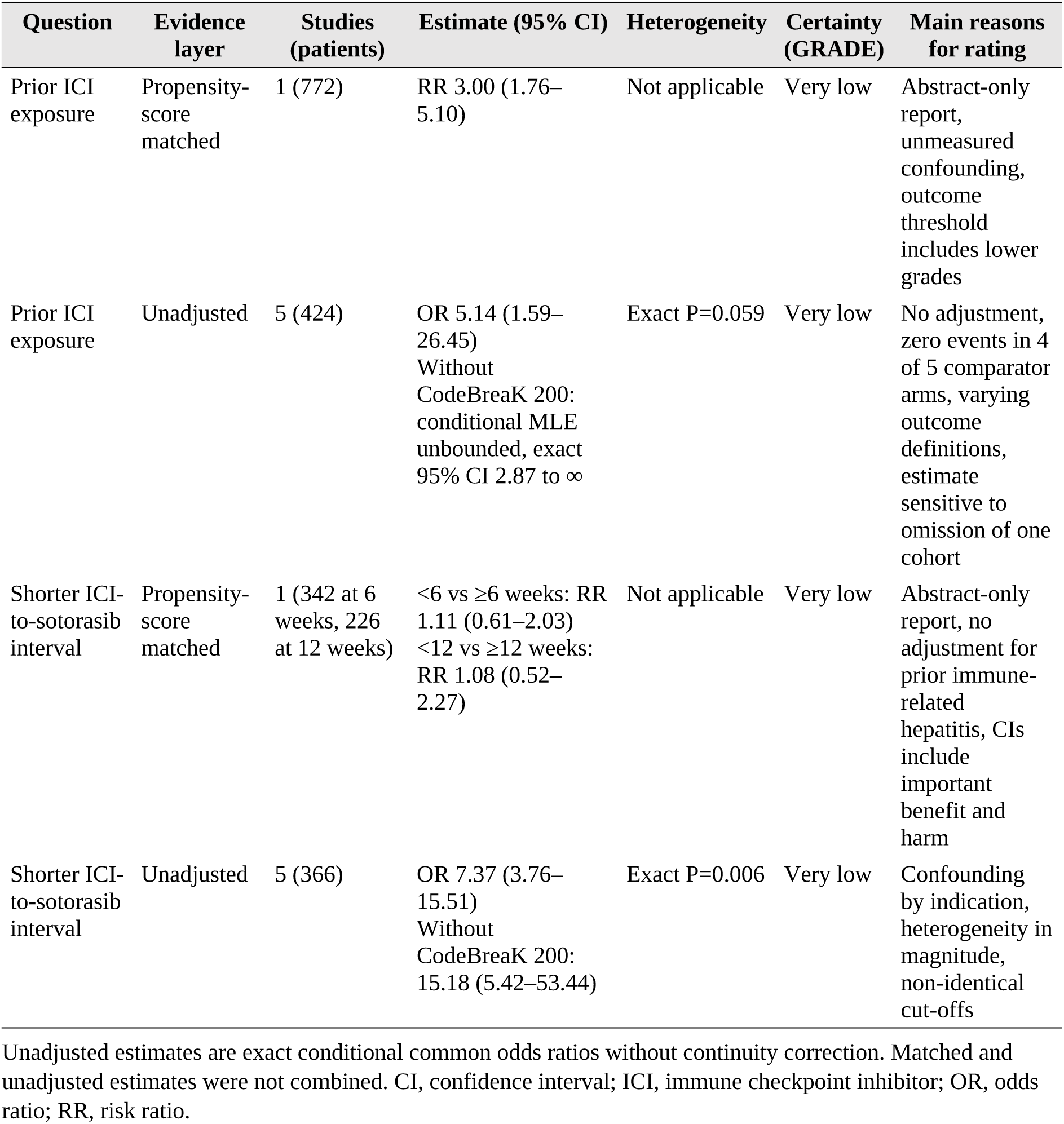
Summary of findings and certainty of evidence.

### Interval between ICI and sotorasib

In the matched analysis, starting sotorasib less than 6 weeks after ICI was not associated with a higher risk than starting at 6 weeks or later (11.7% vs 10.5%, RR 1.11, 95% CI 0.61– 2.03, 171 per arm). The same was true for the 12-week comparison (11.5% vs 10.6%, RR 1.08, 0.52–2.27, 113 per arm) (Figure 3A). Both CIs are compatible with clinically important differences in either direction.

**Figure 3.**
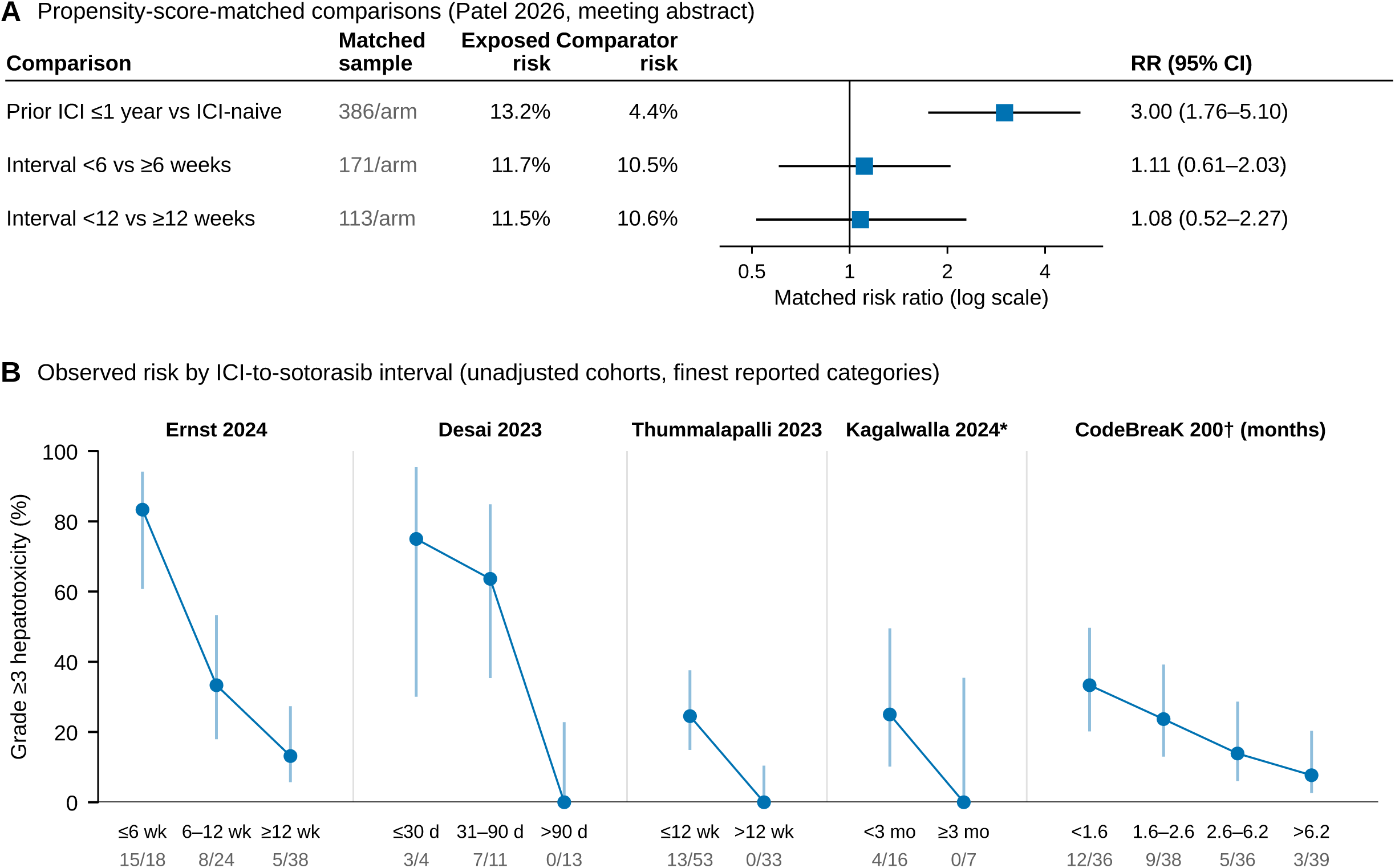
(A) Propensity-score-matched risk ratios (RRs) as reported by Patel et al. (meeting abstract), with observed risks in exposed and comparator groups. (B) Observed risk of grade 3 or higher hepatotoxicity by interval between the last immune checkpoint inhibitor (ICI) dose and sotorasib, in the finest categories reported by each unadjusted cohort. Points show observed proportions with Wilson 95% confidence intervals, and numbers below each category give events/patients. Categories differ between studies and do not represent a common threshold. *Conference abstract. †Appendix Table S16, interval quartile boundaries in months as printed (rounded).

In the unadjusted layer, 71 of 200 patients in shorter-interval groups and 13 of 166 in longer-interval groups had severe hepatotoxicity. The pooled exact conditional OR was 7.37 (95% CI 3.76–15.51, P<0.001) (Figure 2B). The direction was the same in all five cohorts, but the magnitude differed (exact homogeneity P=0.006, Breslow–Day with Tarone correction P=0.092). The OR in CodeBreaK 200 (3.29, 1.28–9.31) was smaller than in the clinical cohorts. Without CodeBreaK 200, the pooled OR was 15.18 (5.42–53.44). In the finest reported categories, risk fell monotonically with longer intervals in every cohort (Figure 3B). In Ernst et al., rates were 83% (15/18), 33% (8/24), and 13% (5/38) at 6 weeks or less, 6 to 12 weeks, and 12 weeks or more.^9^ In CodeBreaK 200, rates were 33% (12/36), 24% (9/38), 14% (5/36), and 8% (3/39) across interval quartiles.^2^

### Separately described comparison

Chour et al. reported severe sotorasib-related hepatotoxicity in 16 of 48 (33%) patients who received anti-PD-(L)1 therapy in the line immediately before sotorasib, compared with 6 of 54 (11%) in a control group in which 87% had received an ICI followed by at least one other regimen.^8^ This comparison was not pooled, because its comparator was not ICI-naive.

### Sensitivity analyses

In leave-one-out analyses of prior ICI exposure, omitting Ernst et al. reduced the OR to 3.30 (95% CI 0.99–17.28). Omitting Desai et al., Thummalapalli et al., or Kagalwalla et al. gave ORs of 4.00 to 4.79 with CIs excluding 1, and omitting CodeBreaK 200 left the conditional maximum likelihood estimate unbounded. For the interval, omitting any single clinical cohort gave ORs from 5.98 to 7.17, all with CIs excluding 1, and omitting CodeBreaK 200 gave 15.18. Excluding the meeting abstract by Kagalwalla et al. gave ORs of 4.79 (1.47– 24.81) and 7.16 (3.63–15.12) (Supplementary Table S3). The conditional random-effects model gave point estimates of 19.09 for prior exposure and 12.74 for a shorter interval, with between-study standard deviations of 1.75 and 0.94 on the log-OR scale. Reliable CIs could not be obtained for this model with five studies. With a half-normal (0.5) prior on the between-study standard deviation, Bayesian posterior medians were 5.57 to 6.44 for prior exposure and 8.22 to 8.89 for a shorter interval under the two reported priors on the mean log-OR. Absolute risk differences are given in Supplementary Table S3.

### Risk of bias and certainty

All sources were at high risk of bias in the confounding domain (Supplementary Table S4). In the clinical cohorts, the interval was determined by clinical circumstances such as disease tempo, recovery from immune-related adverse events, and fitness to start treatment. In CodeBreaK 200, sotorasib was randomized but prior immunotherapy and its timing were not. The matched analysis did not adjust for prior immune-related hepatitis, baseline liver tests, or ICI regimen and did not report balance diagnostics or event counts. Certainty was very low for every question and layer (Table 2). The completed PRISMA 2020 checklist is provided in Supplementary Table S5.

### Evidence not included in the synthesis

In a pharmacovigilance analysis, co-reporting of ICIs with KRAS-targeted drugs was associated with a higher reporting odds ratio for hepatitis (adjusted ROR 3.03, 95% CI 1.49– 5.93).^25^ This design cannot establish sequence, interval, or incidence. In five real-world patients reported by Luo et al., adagrasib given after sotorasib-associated hepatotoxicity was not followed by recurrent treatment-related hepatotoxicity. One of three additional KRYSTAL-1 patients had grade 3 ALT elevation on adagrasib, and these uncontrolled data do not establish comparative drug safety.^26^

## DISCUSSION

This review separates two questions that are often merged. For prior ICI exposure, the evidence points in one direction. The only matched analysis showed a three-fold higher risk, and the unadjusted cohorts, including prospectively collected trial data, showed higher risk after ICI exposure. For the interval between the last ICI dose and sotorasib, the picture is less clear. Every unadjusted cohort had a higher observed event rate at shorter intervals, whereas the matched analysis did not detect a difference at 6 or 12 weeks and could not exclude clinically important effects.

The inclusion of CodeBreaK 200 changed the evidence base. The four clinical cohorts, one prospectively enrolled biomarker cohort and three retrospective series, all had zero severe hepatic events among ICI-naive patients, which left the upper bound of the common OR unidentified. The trial appendix provides CTCAE-graded hepatic event data collected within a phase 3 trial and the only non-zero ICI-naive comparator. Its interval gradient, from 33% in the shortest quartile to 8% in the longest, is less steep than in the clinical cohorts but has the same direction. The difference in magnitude may reflect outcome definition, since the trial counted hepatic events of interest rather than only treatment-related events, and the stronger selection that shapes interval assignment in routine practice.

The discrepancy between the unadjusted interval gradient and the null matched result has several possible explanations. Early initiation after ICI may be a marker of rapidly progressive disease, recent or ongoing immune-related toxicity, or other features that also predict liver injury. Matching can remove only measured confounding, and the matched analysis did not include prior immune-related hepatitis, which was strongly associated with severe hepatotoxicity in the cohort by Ernst et al.^9^ The matched outcome, an aminotransferase increase above three times the upper limit of normal, is not equivalent to the CTCAE grade 3 threshold used by the other studies, so differences in outcome definition could affect the comparison in either direction. The matched timing comparisons were also small, and their CIs do not exclude clinically important effects. The null result therefore does not show that timing is irrelevant, and the unadjusted gradient does not show that delaying sotorasib prevents hepatotoxicity.

Biomarker associations and case reports are compatible with, but do not establish, an ICI-primed mechanism. Ernst et al. observed higher pembrolizumab concentrations in patients with severe hepatotoxicity,^9^ and a biopsy in one published sotorasib case showed a pattern typical of checkpoint inhibitor hepatitis.^4^ Lin et al. reported more hepatotoxicity with crizotinib given after ICI in a comparative cohort,^6^ and Sisi et al. described liver injury with capmatinib after ICI.^7^ A time-varying effect of residual checkpoint blockade is biologically plausible, but the available data neither establish a graded causal relation nor identify a threshold beyond which risk returns to baseline.

These findings have implications for practice. The prescribing information recommends liver tests before sotorasib, every 3 weeks for the first 3 months, and monthly thereafter, and reports any-grade hepatotoxicity in 40% of patients with immunotherapy within 3 months, compared with 18% after a longer interval and 17% without prior immunotherapy.^27^ Prior ICI exposure, particularly when recent, may warrant individualized additional vigilance within this schedule, although the benefit of intensified monitoring has not been tested. The evidence does not establish a mandatory washout, and a fixed delay may be harmful for patients with progressive disease. The decision to start sotorasib soon after ICI should weigh disease tempo, prior immune-related hepatitis, baseline liver function, and the availability of alternative treatments. Whether other KRAS G12C inhibitors carry a similar sequence-dependent risk cannot be determined from the available comparative data.

This review has limitations. All evidence was observational with respect to the exposure, and the only adjusted analysis is a meeting abstract without event counts, balance diagnostics, or a description of its data source. The unadjusted pooled estimates assume a common OR across studies with different outcome definitions and interval cut-offs, and they carry the confounding of their source studies. The interval cut-offs were not identical and should not be read as a common biological threshold. Two sources were meeting abstracts, the matched analysis and a single-centre series of 28 patients, and overlap between the matched analysis and other cohorts cannot be excluded because its data source was not reported. The review was not prospectively registered, and the synthesis approach was finalized after data extraction. Finally, the small number of studies precluded assessment of small-study effects.

Future studies should report patient-level time from the last ICI dose, prior immune-related hepatitis, baseline liver tests, and CTCAE-graded liver events, and should analyze time as a continuous variable with adjustment for disease tempo. Prospective monitoring studies and pharmacokinetic measurement of residual checkpoint antibody would help distinguish an effect of elapsed time from selection.

## CONCLUSIONS

Prior ICI exposure was associated with a higher risk of severe hepatotoxicity with sotorasib in all available comparisons. Shorter intervals between ICI and sotorasib are associated with higher risk in unadjusted data from all available cohorts, including a phase 3 trial, but this association was not confirmed in the only matched analysis. The evidence does not establish a mandatory washout period. Liver-test monitoring as recommended in the prescribing information applies to all patients, additional surveillance specifically after ICI exposure remains an untested precaution, and certainty in all comparative estimates is very low.

## Data Availability

All data were extracted from published reports. The extracted data set, analysis code, search records, and deduplication files are available in the OSF project (https://osf.io/jpm3w/). The review was registered in OSF Registries (doi:10.17605/OSF.IO/X3R9Q).

## ACKNOWLEDGEMENTS

Author contributions: SC: conceptualization, data curation, formal analysis, writing (original draft). HSK: conceptualization, methodology, supervision, funding acquisition, writing (review and editing). Both authors had access to all data, approved the final version, and accept responsibility for the decision to submit.

## DATA SHARING

All data were extracted from published reports. The extracted data set, analysis code, search records, and deduplication files are available in the OSF project (https://osf.io/jpm3w/). The review was retrospectively registered in OSF Registries (doi:10.17605/OSF.IO/X3R9Q).

## ETHICS

This systematic review used only published, aggregate data. Ethics committee approval and informed consent were not required.

## DECLARATION OF GENERATIVE AI AND AI-ASSISTED TECHNOLOGIES IN THE WRITING PROCESS

The authors used Claude (Anthropic) to assist with literature searching, data extraction, statistical programming, and language editing. The authors verified all outputs and take full responsibility for the content.

