## Supplementary Tables S1-S5 for "Prior immune checkpoint inhibitor exposure, treatment interval, and severe hepatotoxicity with sotorasib in KRAS G12C-mutant non-small-cell lung cancer: a systematic review and meta-analysis"

**SUPPLEMENTARY INFORMATION**

**CONTENTS**

**Supplementary Table S1: Search strategies**

**Supplementary Table S2: Reports assessed in full and excluded, with reasons**

**Supplementary Table S3: Sensitivity analyses**

**Supplementary Table S4: Risk of bias (ROBINS-E)**

**Supplementary Table S5: PRISMA 2020 checklist**

**Supplementary Table S1. Search strategies**

**PubMed (searched 26 September 2026, 25 records)**: (sotorasib OR adagrasib OR divarasib OR glecirasib OR garsorasib OR olomorasib OR fulzerasib OR "KRAS G12C inhibitor*") AND (hepat* OR liver OR ALT OR aminotransferase) AND (immunotherap* OR "immune checkpoint" OR pembrolizumab OR nivolumab OR atezolizumab OR durvalumab OR "anti-PD-1" OR "anti-PD-L1" OR sequential)

**Europe PMC (searched 26 September 2026, 407 records; rerun 4 October 2026, 413 records)**: (sotorasib OR adagrasib OR divarasib OR glecirasib OR garsorasib OR olomorasib OR fulzerasib OR "KRAS G12C inhibitor") AND (hepatotoxicity OR "liver injury" OR transaminitis OR hepatitis OR "ALT elevation") AND (immunotherapy OR "checkpoint inhibitor" OR "anti-PD-1" OR "anti-PD-L1" OR pembrolizumab)

**Embase (Embase.com; searched 4–5 October 2026, 92 records)**:

#1 'sotorasib'/exp OR 'adagrasib'/exp OR sotorasib:ti,ab,kw OR adagrasib:ti,ab,kw OR divarasib:ti,ab,kw OR glecirasib:ti,ab,kw OR garsorasib:ti,ab,kw OR olomorasib:ti,ab,kw OR fulzerasib:ti,ab,kw OR 'amg 510':ti,ab OR 'mrtx849':ti,ab OR ('kras g12c' NEAR/3 inhibit*):ti,ab,kw.

#2 'liver toxicity'/exp OR 'drug induced liver injury'/exp OR 'alanine aminotransferase blood level'/exp OR 'aspartate aminotransferase blood level'/exp OR hepatotox*:ti,ab,kw OR 'liver injur*':ti,ab,kw OR transaminit*:ti,ab,kw OR hepatitis:ti,ab,kw OR ((alt OR ast OR aminotransferase* OR transaminase*) NEAR/3 (elevat* OR increas*)):ti,ab.

#3 'immune checkpoint inhibitor'/exp OR 'programmed death 1 receptor antibody'/exp OR 'programmed death 1 ligand 1 antibody'/exp OR immunotherap*:ti,ab,kw OR 'checkpoint inhibit*':ti,ab,kw OR pembrolizumab:ti,ab OR nivolumab:ti,ab OR atezolizumab:ti,ab OR durvalumab:ti,ab OR cemiplimab:ti,ab OR ipilimumab:ti,ab OR 'anti pd 1':ti,ab OR 'anti pd l1':ti,ab OR sequen*:ti,ab.

#4 #1 AND #2 AND #3.

**Web of Science Core Collection (searched 5 October 2026, 20 records)**: TS=(sotorasib OR adagrasib OR divarasib OR glecirasib OR garsorasib OR olomorasib OR fulzerasib OR "AMG 510" OR "MRTX849" OR "KRAS G12C inhibitor*") AND TS=(hepatotox* OR "liver injur*" OR transaminit* OR hepatitis OR "aminotransferase*" OR "transaminase*" OR "liver enzyme*") AND TS=(immunotherap* OR "checkpoint inhibitor*" OR pembrolizumab OR nivolumab OR atezolizumab OR durvalumab OR "anti-PD-1" OR "anti-PD-L1" OR sequen*).

Scopus (searched 5 October 2026, 88 records): TITLE-ABS-KEY((sotorasib OR adagrasib OR divarasib OR glecirasib OR garsorasib OR olomorasib OR fulzerasib OR "AMG 510" OR "MRTX849" OR "KRAS G12C inhibitor*") AND (hepatotox* OR "liver injur*" OR transaminit* OR hepatitis OR aminotransferase* OR transaminase* OR "liver enzyme*") AND (immunotherap* OR "checkpoint inhibitor*" OR pembrolizumab OR nivolumab OR atezolizumab OR durvalumab OR "anti-PD-1" OR "anti-PD-L1" OR sequen*)).

**Cochrane CENTRAL (searched 5 October 2026, 10 records)**: (sotorasib OR adagrasib OR divarasib OR "KRAS G12C"):ti,ab,kw AND (hepatotox* OR liver OR transaminase* OR aminotransferase*):ti,ab,kw.

**Meeting proceedings**: ASCO (Journal of Clinical Oncology supplements), ESMO, ESMO Asia and ELCC (Annals of Oncology and ESMO Open supplements), and IASLC WCLC (Journal of Thoracic Oncology supplements), 2021 to 2026, searched for sotorasib, adagrasib, KRAS G12C, hepatotoxicity, and immunotherapy. Reference lists and trial appendices of included studies were screened.

**Supplementary Table S2. Reports assessed in full and excluded, with reasons**

| **Report** | **Design** | **Reason for exclusion** |
| --- | --- | --- |
| Wislez 2025 (IFCT-2102), Eur J Cancer, doi:10.1016/j.ejca.2025.115301 | Real-world cohort | Outcome not liver-specific: ICI-stratified comparison covered all grade 3–4 TRAEs |
| Passiglia 2024 (Italian EAP), Lung Cancer 187:107444, doi:10.1016/j.lungcan.2023.107444 | Expanded access cohort | Outcome not liver-specific: 23/26 pretreated patients with grade 3–4 TRAEs had prior anti-PD-(L)1; hepatotoxicity not reported by exposure |
| Zhou 2024 (US veterans), JTO Clin Res Rep 5:100670, doi:10.1016/j.jtocrr.2024.100670 | Real-world cohort | Outcome not liver-specific: composite of dose modification for diarrhea, hepatotoxicity, or pneumonitis (26/63 vs 12/65) |
| Skoulidis 2025 (pooled CodeBreaK), Oncologist 30, doi:10.1093/oncolo/oyae356 | Pooled trial analysis | Any-grade hepatotoxicity only (80/240 vs 26/188) |
| Novello 2026 (global EAP), JTO Clin Res Rep 7:100974, doi:10.1016/j.jtocrr.2026.100974 | Expanded access cohort | No stratification by prior ICI |
| Stratmann 2024 (German compassionate use), Eur J Cancer 201:113911, doi:10.1016/j.ejca.2024.113911 | Real-world cohort | Liver enzyme elevation not reported by prior ICI |
| Chour 2024 (two-centre management series), Lung Cancer 191:107789, doi:10.1016/j.lungcan.2024.107789 | Case series | Restricted to patients with grade ≥2 events; no denominator by exposure |
| Dy 2023 (CodeBreaK 100, 2-year), J Clin Oncol 41:3311–3317, doi:10.1200/JCO.22.02524 | Phase 1/2 trial | No extractable counts by prior ICI or interval |
| Luo 2024, JCO Precis Oncol 8:e2300644 | Case series | No ICI-naive comparator; cross-study comparison |
| Li 2024 (FAERS), Cancer Immunol Immunother | Pharmacovigilance | Ineligible design: co-reporting, no exposure timing or incidence |
| Pan 2026 (FAERS), Lung Cancer 214:109311, doi:10.1016/j.lungcan.2026.109311 | Pharmacovigilance | Ineligible design: disproportionality analysis, no prior-ICI comparison |
| Burns 2026 (olomorasib plus pembrolizumab), J Thorac Oncol 21:103528, doi:10.1016/j.jtho.2025.11.018 | Phase 1/2 trial | Concurrent ICI plus KRAS G12C inhibitor, not sequential exposure |
| KRYSTAL-7 first-line adagrasib plus pembrolizumab (meeting abstract LBA65) | Phase 2 trial | Concurrent ICI plus KRAS G12C inhibitor, not sequential exposure |
| Rakshit/Desai, IASLC WCLC 2022 abstract MA13.09 (other methods) | Meeting abstract | Preliminary report overlapping an included cohort (Desai 2023) |

Thirteen reports were identified through database searches and one through other methods. EAP, expanded access programme; FAERS, FDA Adverse Event Reporting System; ICI, immune checkpoint inhibitor; TRAE, treatment-related adverse event.

**Supplementary Table S3. Sensitivity analyses**

| **Analysis** | **Omitted study** | **k** | **Estimate** | **95% CI** | **P** |
| --- | --- | --- | --- | --- | --- |
| Prior ICI (exact conditional OR) | none | 5 | 5.14 | 1.59 to 26.45 | 0.0018 |
| Prior ICI (exact conditional OR) | Ernst | 4 | 3.30 | 0.99 to 17.28 | 0.053 |
| Prior ICI (exact conditional OR) | Desai | 4 | 4.62 | 1.42 to 23.92 | 0.0043 |
| Prior ICI (exact conditional OR) | Thummalapalli | 4 | 4.00 | 1.20 to 20.98 | 0.014 |
| Prior ICI (exact conditional OR) | Kagalwalla | 4 | 4.79 | 1.47 to 24.81 | 0.0042 |
| Prior ICI (exact conditional OR) | CodeBreaK 200 | 4 | NE | 2.87 to ∞ | 0.00016 |
| Shorter interval (exact conditional OR) | none | 5 | 7.37 | 3.76 to 15.51 | 1.2e-11 |
| Shorter interval (exact conditional OR) | Ernst | 4 | 7.17 | 3.13 to 18.63 | 4.9e-08 |
| Shorter interval (exact conditional OR) | Desai | 4 | 5.98 | 3.01 to 12.73 | 7e-09 |
| Shorter interval (exact conditional OR) | Thummalapalli | 4 | 6.36 | 3.16 to 13.66 | 4.4e-09 |
| Shorter interval (exact conditional OR) | Kagalwalla | 4 | 7.16 | 3.63 to 15.12 | 5.3e-11 |
| Shorter interval (exact conditional OR) | CodeBreaK 200 | 4 | 15.18 | 5.42 to 53.44 | 6.1e-11 |
| Prior ICI RD (risk difference, %, Newcombe CI) | Ernst | 1 | 35.0 | 7.4 to 45.9 |  |
| Prior ICI RD (risk difference, %, Newcombe CI) | Desai | 1 | 35.7 | -22.4 to 54.2 |  |
| Prior ICI RD (risk difference, %, Newcombe CI) | Thummalapalli | 1 | 15.1 | -2.8 to 24.2 |  |
| Prior ICI RD (risk difference, %, Newcombe CI) | Kagalwalla | 1 | 17.4 | -27.3 to 37.1 |  |
| Prior ICI RD (risk difference, %, Newcombe CI) | CodeBreaK 200 | 1 | 4.5 | -17.3 to 16.5 |  |
| Shorter interval RD (risk difference, %, Newcombe CI) | Ernst | 1 | 41.6 | 21.1 to 57.5 |  |
| Shorter interval RD (risk difference, %, Newcombe CI) | Desai | 1 | 66.7 | 32.9 to 84.8 |  |
| Shorter interval RD (risk difference, %, Newcombe CI) | Thummalapalli | 1 | 24.5 | 10.4 to 37.6 |  |
| Shorter interval RD (risk difference, %, Newcombe CI) | Kagalwalla | 1 | 25.0 | -13.4 to 49.5 |  |
| Shorter interval RD (risk difference, %, Newcombe CI) | CodeBreaK 200 | 1 | 17.7 | 5.0 to 30.0 |  |
| Prior ICI, conditional random-effects ML | none | 5 | 19.09 | not estimated |  |
| Shorter interval, conditional random-effects ML | none | 5 | 12.74 | not estimated |  |
| Prior ICI, Bayesian RE, τ~HN(0.5), μ~N(0,2.5²) | none | 5 | 5.57 | 1.75 to 26.61 (CrI) |  |
| Prior ICI, Bayesian RE, τ~HN(0.5), μ~N(0,10²) | none | 5 | 6.44 | 1.88 to 39.27 (CrI) |  |
| Shorter interval, Bayesian RE, τ~HN(0.5), μ~N(0,2.5²) | none | 5 | 8.22 | 3.65 to 25.56 (CrI) |  |
| Shorter interval, Bayesian RE, τ~HN(0.5), μ~N(0,10²) | none | 5 | 8.89 | 3.85 to 32.07 (CrI) |  |

Risk differences are exposed minus comparator (shorter minus longer interval). Bayesian results for the other four prior combinations are in the OSF project. CrI, credible interval; HN, half-normal; ML, maximum likelihood; RE, random effects. P values are two-sided exact values based on probability ordering.

**Supplementary Table S4. Risk of bias (ROBINS-E, domain-level judgments)**

| **Study** | **Confounding** | **Exposure measurement** | **Participant selection** | **Post-exposure interventions** | **Missing data** | **Outcome measurement** | **Selection of reported result** |
| --- | --- | --- | --- | --- | --- | --- | --- |
| Ernst 2024 | High | Some concerns | Some concerns | Some concerns | Some concerns | Some concerns | Some concerns |
| Desai 2023 | High | Some concerns | High | Some concerns | Some concerns | Some concerns | High |
| Thummalapalli 2023 | High | Some concerns | Some concerns | High | Some concerns | Some concerns | High |
| Kagalwalla 2024 | High | Some concerns | High | Some concerns | High | Some concerns | High |
| de Langen 2023 | High | Some concerns | High | Some concerns | Some concerns | Some concerns | High |
| Chour 2023 | High | Some concerns | High | Some concerns | Some concerns | Some concerns | High |
| Patel 2026 | High | Some concerns | High | Some concerns | High | High | High |

Judgments are domain-level because most sources did not report enough detail to answer all signalling questions. Supporting quotations for each judgment are provided in the risk-of-bias file in the OSF project.

**Supplementary Table S5. PRISMA 2020 checklist**

| **Item** | **Topic** | **Location in manuscript** |
| --- | --- | --- |
| 1 | Title | Title |
| 2 | Abstract | Abstract |
| 3 | Rationale | Introduction |
| 4 | Objectives | Introduction, final paragraph |
| 5 | Eligibility criteria | Materials and methods, Eligibility criteria |
| 6 | Information sources | Materials and methods, Information sources and search |
| 7 | Search strategy | Supplementary Table S1 |
| 8 | Selection process | Materials and methods, Study selection and data extraction |
| 9 | Data collection process | Materials and methods, Study selection and data extraction |
| 10a–b | Data items | Materials and methods, Study selection and data extraction |
| 11 | Study risk of bias assessment | Materials and methods, Risk of bias and certainty of evidence |
| 12 | Effect measures | Materials and methods, Synthesis methods |
| 13a–f | Synthesis methods | Materials and methods, Synthesis methods |
| 14 | Reporting bias assessment | Materials and methods, Synthesis methods |
| 15 | Certainty assessment | Materials and methods, Risk of bias and certainty of evidence |
| 16a–b | Study selection | Results, Study selection; Figure 1; Supplementary Table S2 |
| 17 | Study characteristics | Results, Study characteristics; Table 1 |
| 18 | Risk of bias in studies | Results, Risk of bias and certainty; Supplementary Table S4 |
| 19 | Results of individual studies | Figures 2 and 3; Table 1 |
| 20a–d | Results of syntheses | Results, Prior ICI exposure and Interval sections; Sensitivity analyses; Figures 2 and 3 |
| 21 | Reporting biases | Materials and methods, Synthesis methods (not assessed statistically) |
| 22 | Certainty of evidence | Table 2; Supplementary Table S3 |
| 23a–d | Discussion | Discussion |
| 24a–c | Registration and protocol | Materials and methods, Protocol and reporting; Data sharing |
| 25 | Support | Funding |
| 26 | Competing interests | Disclosure |
| 27 | Availability of data, code, and other materials | Data sharing |

Item numbers follow Page et al., BMJ 2021;372:n71.
